# Copy-number variants and polygenic risk for intelligence confer risk for autism spectrum disorder irrespective of their effects on cognitive ability

**DOI:** 10.1101/2023.11.29.23299190

**Authors:** Zoe Schmilovich, Vincent-Raphaël Bourque, Elise Douard, Guillaume Huguet, Cécile Poulain, Jay P. Ross, Paria Alipour, Charles-Étienne Castonguay, Nadine Younis, Martineau Jean-Louis, Zohra Saci, Zdenka Pausova, Tomas Paus, Gunter Schuman, David Porteous, Gail Davies, Paul Redmond, Sarah E. Harris, Ian J. Deary, Heather Whalley, Caroline Hayward, Patrick A. Dion, Sébastien Jacquemont, Guy A. Rouleau

**Author notes:** **Corresponding Author:** Dr. Guy A. Rouleau, MD, Ph.D., FRCPC, Department of Neurology and Neurosurgery, McGill University, Montréal, Québec, Canada, H3A 2B4.

## Abstract

Rare copy number variants (CNVs) and polygenic risk for intelligence (PRS-IQ) both confer risk for autism spectrum disorder (ASD) but have opposing effects on cognitive ability. The field has struggled to disentangle the effects of these two classes of genomic variants on cognitive ability from their effects on ASD risk, in part because previous studies did not include controls with cognitive measures. We aim to investigate the impact of these genomic variants on ASD risk while adjusting for their known effects on cognitive ability. In a cohort of 8,426 subjects with ASD and 169,804 controls with cognitive assessments, we found that rare coding CNVs and PRS-IQ increased ASD risk, even after adjusting for their effects on cognitive ability. Bottom decile PRS-IQ and CNVs both decreased cognitive ability but had opposing effects on ASD risk. Models combining both classes of variants showed that the effects of rare CNVs and PRS-IQ on ASD risk and cognitive ability were largely additive, further suggesting that risk for ASD is conferred independently from its effects on cognitive ability. Despite imparting mostly additive effects on ASD risk, rare CNVs and PRS-IQ showed opposing effects on core and associated features and developmental history among subjects with ASD. Our findings suggest that cognitive ability itself may not be the factor driving the underlying risk for ASD conferred by these two classes of genomic variants. In other words, ASD risk and cognitive ability may be two distinct manifestations of CNVs and PRS-IQ. This study also highlights the challenge of understanding how genetic risk for ASD maps onto its dimensional traits.

## INTRODUCTION

ASD has a high estimated overall heritability and a genetic architecture composed of rare and common variants^1–4^. Despite its strong genetic component, there remains insufficient evidence for ASD-specific genes^5^. Rare *de novo* and inherited copy-number variants (CNVs) that substantially increase ASD risk are present in 8-14% of individuals with ASD^6,7(p20)^. These ASD-susceptibility variants also have negative effects on cognition^3^.

Studies have quantified the negative effects of CNVs on cognitive ability, showing that the effects are the same in individuals with ASD and the general population^8, 9^. Despite this, studies have also shown that deletion and duplication CNVs increase the risk for ASD even after adjusting for their effects on cognitive ability^10^. In other words, CNVs are over-represented in cases with ASD compared to controls with similar cognitive capacities. Conversely, there is a significant genetic correlation (r_G_ = 0.199) between ASD and intelligence such that common variants that increase the risk for ASD also increase cognitive ability in the general population^2^. Studies also suggest that PRS-ASD is positively associated with intelligence and higher educational attainment^11, 12^. This effect is opposite to what has been shown for PRSs of other neurodevelopmental disorders^13–15^. These findings are counterintuitive and difficult to interpret, given that individuals with ASD have IQ levels that are on average 1 standard deviation (s.d.) below the general population^16^. Indeed, there is a complex relationship between genetic risk, cognitive ability, and ASD that is paradoxical and remains contentious.

Previous studies have estimated the effects of rare and common variants in different samples separately; as such, combined variant effects on ASD risk and cognitive ability have not been assessed, nor have their effect on other ASD-associated traits been evaluated. This study aims to clarify the individual and combined impact of rare variants that decrease cognitive ability (CNVs) and common variants that increase intelligence (PRS-IQ) on ASD risk, cognitive ability, and other ASD-associated traits.

## METHODS

### Datasets

Three ASD cohorts and five general population cohorts were included in the study (**Table ST1**).

#### ASD cohorts

Three family-based ASD cohorts were included in the study: the Simons Simplex Collection (SSC)^17^, Simons Foundation Powering Autism Research for Knowledge (SPARK)^18^, and MSSNG^19^. The SSC and SPARK cohorts comprised SNP genotyping data, while the MSSNG cohort comprised Whole-Genome Sequencing (WGS) data. In total, the genetic data of 28,307 cases with ASD and 50,953 typically developing family members (including siblings and parents of the affected proband(s)) were included in this study. The unaffected family members from the ASD cohorts were used as intrafamilial controls (n=50,953) in the study.

#### Unselected general population cohorts

Five community-based cohorts were used as extrafamilial controls: IMAGEN^20^; Generation Scotland (GS)^21^; Lothian Birth Cohort 1936 (LBC)^22, 23^; the Saguenay Youth Study (SYS)^24^, and the UK BioBank (UKBB) cohort^25^. To avoid base and target sample overlap in PRS computation, we excluded the UKBB subjects that were part of the 2017 intelligence GWAS (150,000 subjects from the May 2015 release of the genotype data^15^). As such, this study only includes the imputed genotyping data from the second data release (July 2017) of the UKBB cohort. In total, the genetic data of 357,546 extrafamilial controls were included in this study.

### CNV calling, filtering, and annotation

CNVs were detected from the genotyping and WGS data across all eight cohorts and filtered according to established methods (https://martineaujeanlouis.github.io/MIND-GENESPARALLELCNV/)^8–10^. In brief, in order to minimize the number of false discoveries, the pipeline involves two algorithms, PennCNV^26^ and QuantiSNP^27^ of which it extracts consensus results. Coding genes fully encompassed in deletion or duplication CNVs were annotated according to two haploinsufficiency scores based on previous observations^10^: the probability of being loss-of-function intolerant (pLI)^28^ and the loss-of-function observed/expected upper bound fraction (LOEUF)^29^ constraint score. In total, the CNVs that were called encompassed 19,368 genes genome-wide, 16,967 encompassed by deletions and 19,282 by duplications. Of these genes, 18,347 (94.7%) had available pLI and LOEUF annotation data available; 16,047 (94.6%) for deletions and 18,264 (94.7%) for duplications.

While both scores reflect genetic fitness, they were used in different contexts in this study, and their distinct distribution of intolerance scores across genes reflects the nature of how they were derived. The pLI score (ranges from 0 to 1, from most to least tolerant) was designed to capture high-confidence genes harbouring protein truncating variants (PTVs)^30^ and is generally used as a dichotomous metric. We observed that the pLI score had a bimodal distribution across the 19,197 genes included in the analysis (**Figure S1a**). This observation suggests that a sum of pLI scores of impacted genes is well suited to capture the genome-wide burden of CNVs per individual (CNV risk score), with higher values denoting a higher burden. On the other hand, the LOEUF scores presented a continuous distribution (ranging from 0.03 to 2) thus better suited to capture the moderate haploinsufficiency of most genes (**Figure S1b),** with lower strata denoting high intolerance to loss of function. Accordingly, a sum of pLI scores of impacted genes was used as a measure of burden per individual. In contrast, the LOEUF annotation was used to stratify individuals according to the number of deleted or duplicated genes intolerant to loss-of-function that they carried. Hereafter, we use the term “intolerant CNVs” to refer to CNVs that encompass constraint genes that are intolerant to loss-of-function.

### Copy-number variant risk score

For each individual, pLI scores for genes fully encompassed within CNVs were summed for deletions (∑*DEL pLI*) and duplications (∑*DUP pLI*) separately. The sum of pLI represents the individual-level burden of deletions and duplications and is referred to as the CNV risk score.

### Genetic quality control and imputation

Genotype and sample-level quality control (QC) were performed across cohorts separately using established criteria ^31^ and the PLINK toolset (v1.9)^32^. The PLINK files for the MSSNG WGS pVCF data were generated using the standard protocol based on the UK BioBank WGS^33^. We used bcftools (v1.13) to remove indels, keep biallelic sites, and normalize the SNV dataset^34^. Then, we used the PLINK toolset (v1.9) to convert the pVCF to the genotyping format data (.bed, .bim, .fam) while applying the following filters: --geno 0.05, --mind 0.05, --maf 0.01 and --hwe 5e8.

In brief, we excluded individuals with genotyping rate <95%, excessive heterozygosity (± 3 standard deviations from the mean using the —indep-pairwise command with a 50-variant window and pruning variants with r^2^ > 0.2), sample missingness >0.02, mismatched in reported and genetic sex, and families with Mendelian errors >5%. We removed SNPs with a call rate <98%, a minor allele frequency (MAF) <1%, deviation from Hardy-Weinberg Equilibrium (HWE) (P <1*x*10^-6^), had Mendelian errors in more than 10% of the families, and SNPs that were not genotyped in more than 10% of families. Population stratification and inference were performed using the Kinship-based INference for Genome-wide association studies (KING) toolset^35^. Given the Eurocentric genome-wide association study (GWAS) summary statistics available, only individuals with ≥85% probability of inferred European ancestry were selected. The filtered genotyping data was imputed on the 1000 Genomes Phase 3 reference panel using the Sanger Imputation Server (https://www.sanger.ac.uk/tool/sanger-imputation-service/) and EAGLE+PBWT pipeline. Given that the number of variants in the MSSNG cohort (>7M), generated from WGS data, was comparable to the number of SNPs in the genotyping datasets following imputation, the MSSNG WGS data were not imputed. For the UKBB cohort, we obtained the imputed genotyping data and selected the samples with self-reported white British or European ancestry that had a genotyping call rate >98%.

The imputed genotypes across all cohorts and technologies were merged, such that only variants that were present across all technologies were retained. Established QC filtering criteria were applied to exclude variants that had poor imputation scores (INFO ≥ 0.3); non-biallelic; MAF <5%; call rate <98%, and deviated from Hardy-Weinberg Equilibrium (P < 5×10-7).

### Polygenic risk score (PRS) generation

Details of the methodological pipeline to compute PRS are detailed in **Figure S2.** To avoid target and base sample overlap, the largest GWAS summary statistics available for intelligence that excluded most of the samples included in the study, were used to compute the polygenic risk score for intelligence (PRS-IQ)^15^. The Sniekers *et al.* (2017) GWAS (n=78,308) used Spearman’s g or a primary measure of fluid intelligence as the outcome for the association. With the exception of the LBC cohort, none of the cohorts used in this study were included in the Sniekers *et al.* GWAS. PRS-CS was used with its default parameters to infer the posterior effect sizes of SNPs in the samples that overlapped with the selected GWAS summary statistics and linkage disequilibrium (LD) 1000 Genomes European reference panel^36^. The PLINK 1.9 “score” parameter was used to estimate the individual-level burden of all scored variants into a PRS for intelligence (PRS-IQ).

To account for subtle population structure differences amongst the European samples, PRS-IQ was modelled as a function of the top 20 ancestry principal components (PCs) in a linear model as follows:

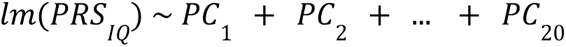

The residuals from the model were extracted to represent a PRS-IQ that removes the underlying effects of ancestry. The PRS-IQ was then scaled according to the mean PRS-IQ of the community-based general population cohorts (UKBB, GS, IMAGEN, LBC, SYS; n = 311,811). Thus, PRS-IQ is represented as the number of standard deviations that an individual is from the unselected general population mean PRS-IQ. The distribution of PRS-IQ across each analysis group and cohort is shown in **Figure S3**.

### Phenotypic measures

Cognitive ability was assessed based on non-verbal IQ (IMAGEN, SYS-children, SSC, MSSNG), parent-report full-scale IQ or non-verbal IQ (SPARK), g-factor (GS, SYS-parents, LBC), or both g-factor and fluid intelligence (FI) (UKBB). For a detailed description of the evaluation of IQ across cohorts see Huguet *et al.* (2021)^9^. The g-factor represents the first dimension obtained by principal component analysis of cognitive tests primarily assessing fluid reasoning. In SPARK, parent-report full-scale IQ was reported in the form of 10 IQ bins; each bin was assigned the median value from the distribution of non-verbal IQ extracted from charts in a subset of the same cohort. For UKBB, the g-factor was computed using four cognitive tasks assessed in person (N=73,882) and online (N=62,080): Trail Making Test parts A and B (Executive function), Symbol Digit Substitution Test (Processing speed), Paired Associate Learning Test (Verbal declarative memory) and Picture Vocabulary (Crystallized ability). In UKBB, the FI score was assessed in person (N=88,441) and online (N=13,773).

The NVIQ and g-factor measures across the cohorts were standardized to reflect the general intelligence of the samples based on cognitive tests that primarily assessed cognitive ability ^9^. All cognitive ability measures were z-scored within each cohort and residualized for age using a linear regression as detailed in Huguet et al. (2021). The distribution of cognitive ability is shown in **Figure S4**.

We used a linear regression model to assess the mean change in cognitive ability as a function of ASD case/control status while adjusting for sex as a covariate:

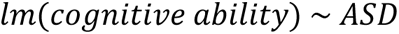

To further detail the clinical heterogeneity among ASD subjects, we selected a range of phenotypes (**Figure S5**) that represent fundamental dimensions associated with ASD: 1–Core ASD features: Repetitive Behavior Scale-Revised (RBSR^37^), Social Responsiveness Scale parent report (SRS^38^), Social Communication Questionnaire lifetime version (SCQ^39^); 2–ASD specifiers and associated features: Vineland Adaptive Behavior Scale (VABS) composite score and subscales^40^), Developmental Coordination Disorder Questionnaire (DCDQ)^41^), and 3–Developmental history, as per parent report: age at first word, age at first phrase, age at walking independently, language regression.

### Statistical analyses

All statistical analyses were performed using R version 4.0.5^42^.

#### Analysis groups

All statistical analyses were conducted across four case-control groups: 1–cases with ASD (n = 21,255) vs. unselected general population (extrafamilial controls) (n = 311,811); 2–cases with ASD (n = 8,426) vs. unselected general population (extrafamilial controls) (n = 169,804) that had cognitive data available; 3–cases with ASD (n = 21,255) vs. their unaffected family members (intrafamilial controls) (n = 24,474), and; 4–unaffected family members (treated as cases) (n = 24,474) vs. unselected general population controls (n = 311,811). The effect of the genetic factors on cognitive ability was modelled separately in cases with ASD and extrafamilial controls. When comparing subjects with ASD to their unaffected family members, familial relationship was included as a random effect variable. All regression models included sex (and when available, cognitive ability) as a covariate.

#### Impact of CNV burden and PRS-IQ on ASD risk and cognitive ability

To estimate the individual and interactive effect between the sum of deletions (∑DEL_pLI_) and duplications (∑DUP_pLI_) with PRS-IQ on ASD risk, we used a logistic regression model using the *glm* function from “stats” base R package^42^ as follows:

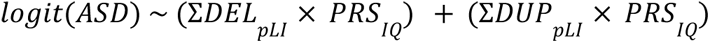

where a binary diagnosis of ASD is the outcome, ∑*DEL*_*pLI*_ and ∑*DUP*^*pLI*^ represent the global burden of deletions and duplications that interact with PRS-IQ.

For analyses comparing cases with ASD to their unaffected family members (intrafamilial controls), a generalized linear mixed-effects (GLME) model was applied using the *glmer* function from the “lme4*”* R package^43^. This model accounted for the effects of relatedness among ASD individuals and the intrafamilial controls by including the family identifier as a random effect.

We also ran a linear regression model with the same predictor variables and cognitive ability as the outcome using the *glm* function from “stats” base R package^42^ as follows:

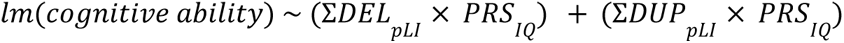

where cognitive ability is a continuous measure standardized to reflect general intelligence as described above. This regression model was computed separately for cases with ASD (n = 8,426) and extrafamilial general population controls (n = 169,804) separately.

All *P* values were adjusted by the Benjamini–Hochberg false-discovery rate (FDR) correction for multiple comparisons using the *p.adjust* function from the base R package.

#### Sliding window analyses across PRS-IQ deciles

We also examined the effect of each PRS-IQ decile on ASD risk and cognitive ability. To do this, we ran ten logistic or linear regressions as follows (for each PRS-IQ decile), depending on whether the outcome was a binary ASD diagnosis or cognitive ability, respectively:

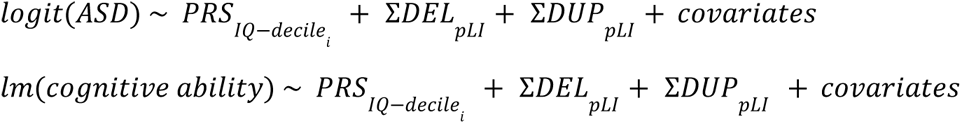

Each model included sex as a covariate, cognitive ability in the logistic regression model when available, and family identifier as a random effect variable when modelling cases with ASD versus intrafamilial controls.

#### Role of deletions and duplications encompassing highly intolerant genes in modulating the impact of PRS-IQ on ASD risk and cognitive ability

To further investigate how CNVs in highly intolerant genes may modulate the impact of PRS-IQ on ASD risk and cognitive ability, we stratified all subjects with available cognitive data into three groups: 1–subjects carrying CNVs in two or more highly constrained genes; 2–subjects carrying a CNV in one highly constrained gene, and; 3–subjects that carried no CNVs in highly constrained genes. A highly constrained gene was defined as having a LOEUF score ≥0.35^29^. This stratification according to CNV carrier status was performed for deletions and duplications separately. The number of cases with ASD and extrafamilial controls in each CNV carrier category is detailed in Table ST4.

We then estimated the impact of PRS-IQ on ASD risk and cognitive ability using the CNV carrier status of the individuals (i.e.: carrying none, one, or two or more deletions or duplications in haploinsufficient genes) as a moderator variable as follows:

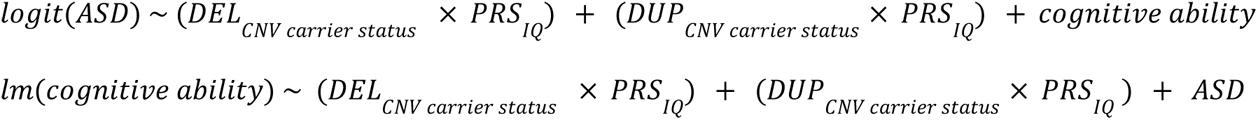

Cognitive ability and ASD case status were used as covariates in the logistic and linear regression models, respectively. Both models included sex as a covariate.

The *sim_slopes* function from the “interactions” R package was used to extract the effect of PRS-IQ on ASD risk and cognitive ability for the three CNV carrier status outcomes. The results were visualized using the *interaction_plot* function from the “interactions” R package for the two ASD risk and cognitive ability models. PRS-IQ and CNV carrier status were specified as the predictor and moderator variables, respectively. The impact of PRS-IQ on ASD risk and cognitive ability, as moderated by CNV carrier status, was performed for deletions and duplications separately.

To further understand the relationship between CNV carrier status, PRS-IQ, and cognitive ability, we ran the following regressions that explored the three-way interaction between the variables:

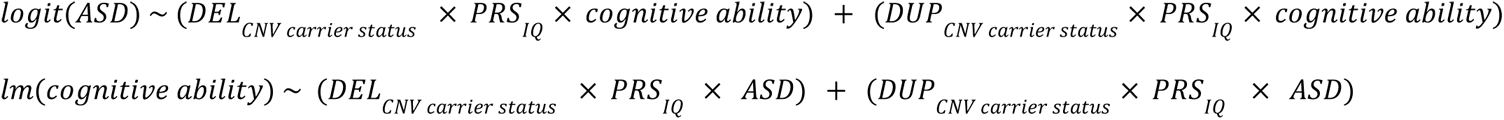

Here, the logistic and linear regression models used cognitive ability and ASD diagnostic status as secondary moderator variables, respectively. Both models included sex as a covariate.

The logistic regression model assessed the interaction between PRS-IQ and CNV carrier status on ASD risk depending on the level of cognitive ability of the subject. Using PRS-IQ as the predictor and CNV carrier status as the moderator, the *sim_slopes* function used cognitive ability as its second moderator. As cognitive ability is a continuous variable, the estimate of PRS-IQ (slope of the focal predictor) on ASD risk for each CNV carrier status was determined according to the mean and ±1 standard deviation values of this moderator. For the linear regression, the estimate of PRS-IQ on cognitive ability for each CNV carrier status was evaluated across both levels of the ASD status moderator (i.e.: case or extrafamilial control) separately. These interactions were visualized using the *interact_plot* function from the “interactions” R package.

#### Convergence of CNV burden and PRS-IQ effect sizes on phenotypic measures among cases with ASD

The distributions of all continuous traits were visualized for normality, after which some variables were applied a logarithmic transformation to normalize their distribution (age of first word, age of first phrase, age of walking independently, and RBSR subscales). All continuous variables were then z-scored before analyses.

Regarding ASD as the outcome, we focused on comparing cases with ASD versus extrafamilial controls. Regarding cognitive ability and other phenotypes, we focused on analyses restricted to cases with ASD.

Linear models for continuous traits and logistic models for binary traits were estimated for each variable as the outcome as follows:

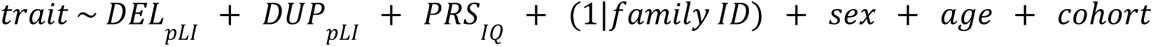

We included a random effect for the family identifier to account for relatedness, and we corrected for cohort where applicable. For ASD and cognitive ability, regressions were adjusted for sex. For other traits, the inclusion of covariates sex and age, where applicable, was based on Akaike’s information criterion (AIC). *P* values were adjusted with the false discovery rate (FDR) method considering the total number of models tested. For all analyses, the significance threshold was fixed at a *FDR*<0.05, two-sided.

## RESULTS

Following quality control, 21,255 cases with ASD; 24,474 unaffected family members of ASD probands (intrafamilial controls), and; 311,811 unselected individuals from the general population (extrafamilial controls) were included in the analyses. Of these, 8,426 cases and 169,804 extrafamilial controls had cognitive data available (**Table 1**). On average, cases with ASD had a 0.69 (95% confidence interval (CI) = [-0.71, -0.67]) lower scaled cognitive ability compared to extrafamilial controls (**<u>Figure S4a</u>**).

**Table 1.**
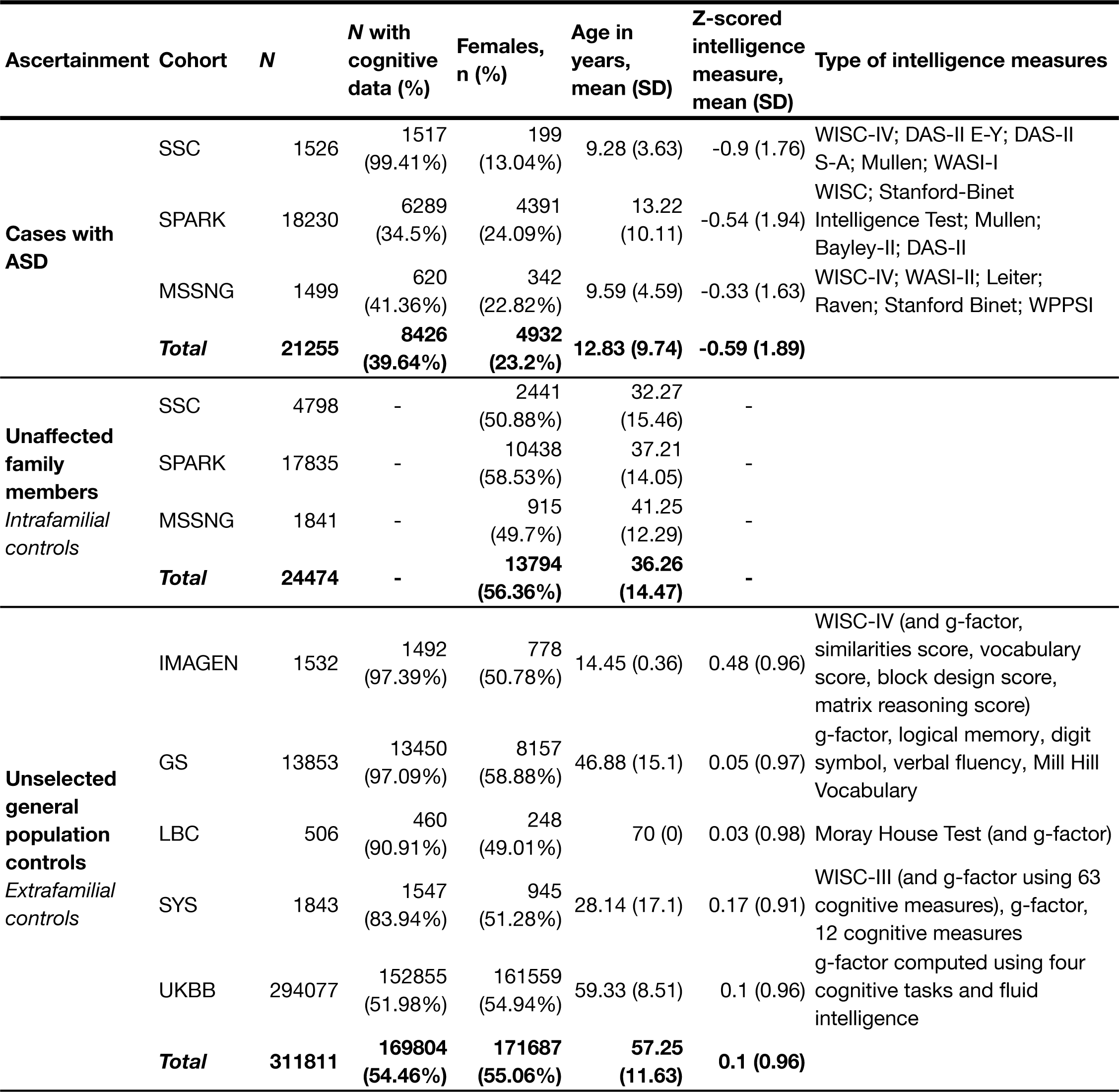
Cohort descriptives following sample and genetic-level quality control. Following quality control, this study uses the genetic data of 357,540 individuals across eight cohorts. These samples include 21,255 individuals diagnosed with ASD, 24,474 unaffected family members from simplex and multiplex families with ASD and 311,811 unselected individuals from community-based general population cohorts. To avoid base and target sample overlap for PRS computation, we only included the UKBB participants from the second data release. Only the subset of individuals with available cognitive data was included in analyses where cognitive ability was included in the regression models.

### Effects of intolerant genes on liability for ASD and cognitive ability when deleted or duplicated

We previously showed that genome-wide deletions and duplications measured by pLI are associated with ASD risk and decreased cognitive ability^8–10^. We sought to extend this analysis to a larger dataset and found that the burden of intolerant genes increased the risk for ASD when deleted or duplicated (OR_DEL_=1.49 [1.45, 1.54]; OR_DUP_=1.18 [1.16, 1.20]) and decreased cognitive ability with similar effect size in cases with ASD (β_DEL_=-0.16 [-0.21, -0.11]; β_DUP_=-0.09 [-0.14, -0.05]) and extrafamilial controls (β_DEL_=-0.11 [-0.13, -0.09]; β_DUP_=-0.06 [-0.07, -0.05]) (Figure 1a, Table ST2).

**Figure 1.**
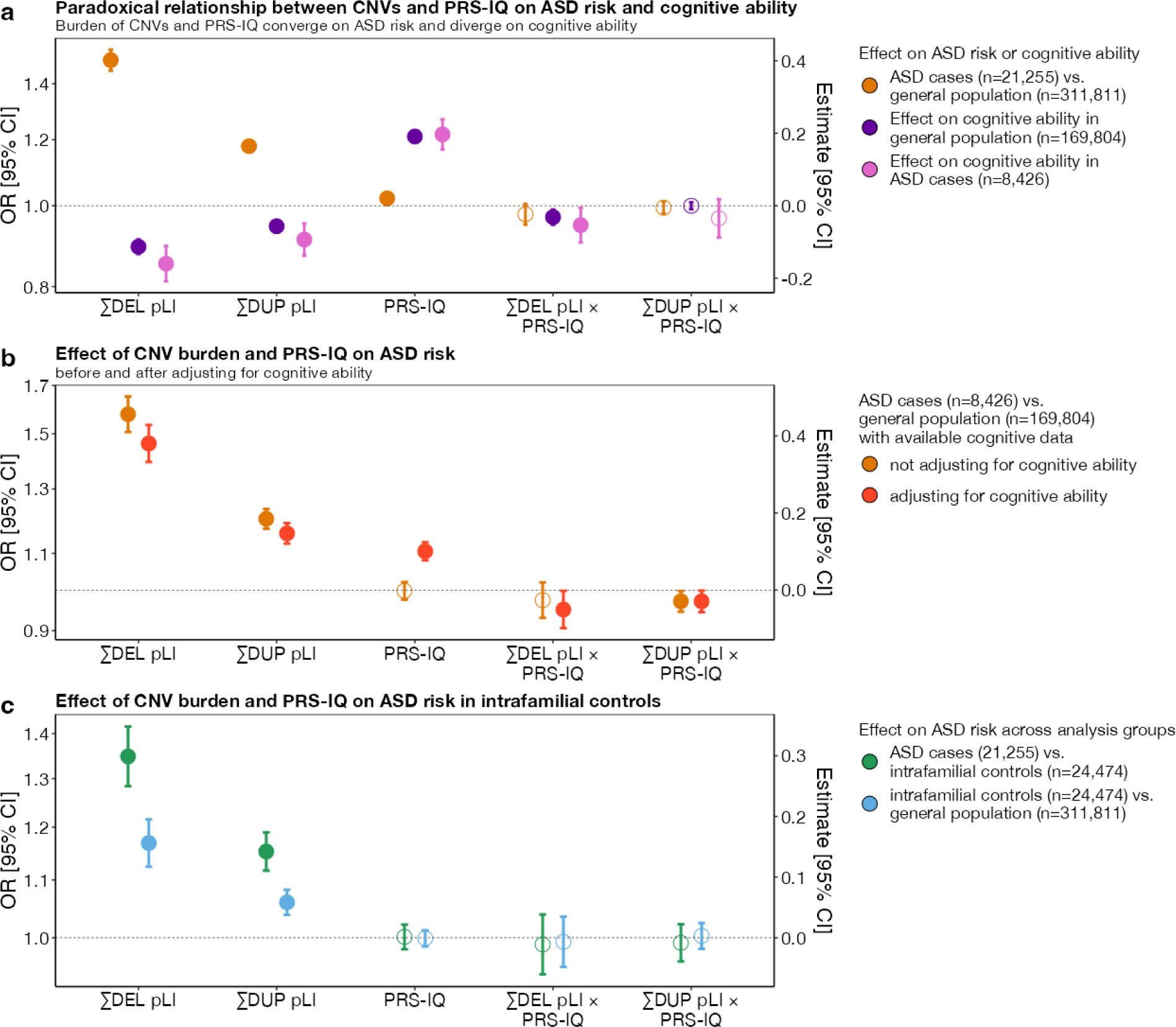
The impact of CNV burden and PRS-IQ on ASD risk and cognitive ability. The estimate and 95% confidence interval (CI) of genetic factors related to cognition in conferring risk for ASD and impact on cognitive ability. **a**) A greater burden of CNV deletions and duplications increase the risk for ASD and decrease cognitive ability (divergent effects). Conversely, PRS-IQ increases the risk for ASD and increases cognitive ability (convergent effects). **b**) Adjusting for cognitive ability does not change the risk for ASD conferred by CNVs and PRS-IQ. The risk for ASD is evaluated only in a subgroup of cases with ASD (n=8,426) and extrafamilial controls (n**=**169,804) for which cognitive ability data were available. CNVs and PRS-IQ increase the risk for ASD, independently from their effects on cognitive ability. **c**) The impact of deletions and duplications on ASD risk is significant – albeit, lower – when comparing subjects with ASD to their unaffected family members versus extrafamilial controls. Although they do not have a diagnosis of ASD, intrafamilial controls have an excess burden of deletions and duplications in comparison to the general population. A similar PRS-IQ between intrafamilial and extrafamilial controls suggests that the differences in PRS-IQ in **a** and **b** are not driven by batch effects between ASD and general population cohorts. Filled-in points represent statistically significant terms (*P* value ≤ 0.05 following FDR adjustment for multiple corrections). Error bars represent the 95% CIs. For detailed model results, see **Table ST2.**

Liability for ASD remained unchanged for deletions and duplications of intolerant genes even after adjusting for their effects on cognitive ability (OR_DEL_=1.46 [1.39, 1.53]; OR_DUP_=1.16 [1.13, 1.19]) (**Figure 1b, Table ST2**).

Because most of the genetic risk examined above is inherited from unaffected parents, we also assessed the burden of risk variants in unaffected ASD family members. While cases with ASD carried a greater burden of deletions and duplications compared to their unaffected family members (OR_DEL_=1.34 [1.28, 1.42]; OR_DUP_=1.15 [1.12, 1.19]) (**Figure 1c, Table ST2**), we found that unaffected family members still carried a greater burden compared to extrafamilial controls (OR_DEL_=1.17 [1.12, 1.22]; OR_DUP_=1.06 [1.04, 1.08]) (**Figure 1c, Table ST2**).

### High PRS-IQ confers risk, and low PRS-IQ decreases the risk for ASD when adjusting for cognitive ability

The positive correlation between PRS-IQ and ASD risk remains misunderstood. To further investigate this relationship, we compared PRS-IQ in subjects with ASD to control participants while accounting for CNV burden and cognitive abilities.

We found that individuals with ASD had a greater PRS-IQ (OR=1.02 [1.00, 1.03]) (**Figure 1a, Table ST3**), despite having a mean score of cognitive ability that was 0.69 (z-score) lower than the extrafamilial controls (β=-0.69 [-0.71, -0.67]) (**Figure S4a**). This increase was more pronounced when adjusting for cognitive ability (**Figure 1b, Table ST2)** (OR=1.10 [1.08, 1.13]). Sensitivity analyses showed that there were no differences in PRS-IQ between intrafamilial controls and controls from general population cohorts (**Figure 1c**).

To assess the specific effects of PRS-IQ on liability for ASD and cognitive ability, and to test for the presence of non-linear effects, we computed effect sizes for each PRS-IQ decile on liability for ASD (**Figure 2a**) and cognitive ability (**Figure 2b**). The top and bottom deciles of PRS-IQ significantly increased and decreased the risk for ASD, respectively, even after adjusting for the effects of cognitive ability. The effects of PRS-IQ on cognitive ability followed a similar pattern to its effects on ASD risk. Furthermore, the impact of PRS-IQ on cognitive ability was identical among subjects with ASD and extrafamilial controls.

**Figure 2.**
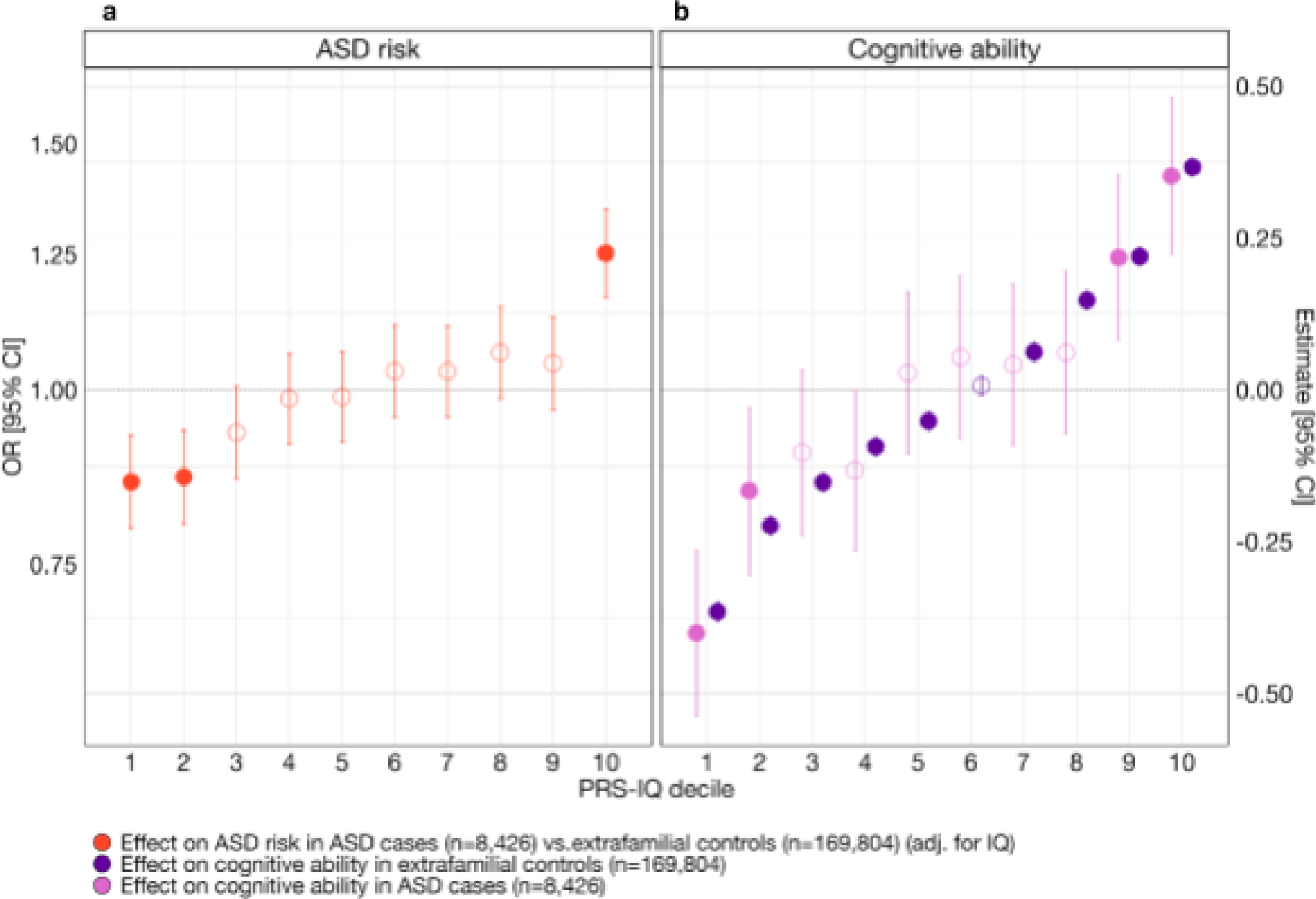
The effect of PRS-IQ deciles on ASD risk and cognitive ability. The estimate and 95% CI of each PRS-IQ decile on ASD risk and cognitive ability. Each regression accounted for the individual-level burden of deletions (∑DEL_pLI_), duplications (∑DUP_pLI_), and – when available – cognitive ability of individuals included in the model. **a**) Even after adjusting for the effects of cognitive ability, a high PRS-IQ (10th decile) increases the risk for ASD, while a low PRS-IQ (1st, 2nd deciles) decreases the risk for ASD. **b**) A PRS-IQ below and above the 6th decile significantly decreases and increases cognitive ability in the general population, respectively. The effect of PRS-IQ on cognitive ability is the same in cases with ASD and in the general population (extrafamilial controls). Filled-in points represent statistically significant terms (*P* value ≤ 0.05 following FDR adjustment for multiple corrections). Error bars represent the 95% CIs. For detailed model results, see Table ST3. See **Figure S6** and **Table ST3** for the results across other analysis groups.

### Deletions and duplications in highly intolerant genes modulate the impact of PRS-IQ on ASD risk and cognitive ability

We identified the same negative interaction between deletions and PRS-IQ on cognitive ability in individuals with ASD (β_DEL*PRS-IQ_=-0.05 [-0.10, -0.006]) and in extrafamilial controls (β_DEL*PRS-IQ_=-0.032 [-0.05, -0.01]) (**Figure 1a**). This suggests that the negative impact of deletions on cognitive ability may be attenuated in individuals with an increasing PRS-IQ, regardless of a diagnosis of ASD.

We also observed a negative interaction between deletions and PRS-IQ, as well as duplications and PRS-IQ, in conferring liability for ASD when adjusting for cognitive ability (OR_DEL*PRS-IQ_= 0.95 [0.91, 1.00]; OR_DUP*PRS-IQ_=0.97 [0.94, 1.0]) (**Figure 1b**). This similarly suggests that the effect of deletions and duplications on ASD risk is reduced in individuals with a high PRS-IQ.

To further examine the relationship between CNVs, PRS-IQ, cognitive ability, and ASD risk, we stratified individuals according to whether they carried zero, one, two or more deletions or duplications in a highly intolerant gene (LOEUF≤0.35) (Table ST4). Then, we assessed the effect of PRS-IQ on ASD risk (**Figures 3a, b**) and cognitive ability (**Figures 3e, f**) using their CNV carrier status (i.e.: the number of deleted or duplicated highly intolerant genes) as a moderator variable. The results suggest that in the absence of deleted or duplicated highly intolerant genes, PRS-IQ has a positive linear impact on ASD risk and cognitive ability. Carrying one deletion in a highly intolerant gene attenuates the impact of PRS-IQ on ASD risk and cognitive ability. In the context of deletions and duplications of two or more intolerant genes, we no longer observed the effect of PRS-IQ on ASD risk and cognitive ability. The impact of PRS-IQ on ASD risk and cognitive ability was similarly attenuated among carriers of duplications in highly intolerant genes.

**Figure 3.**
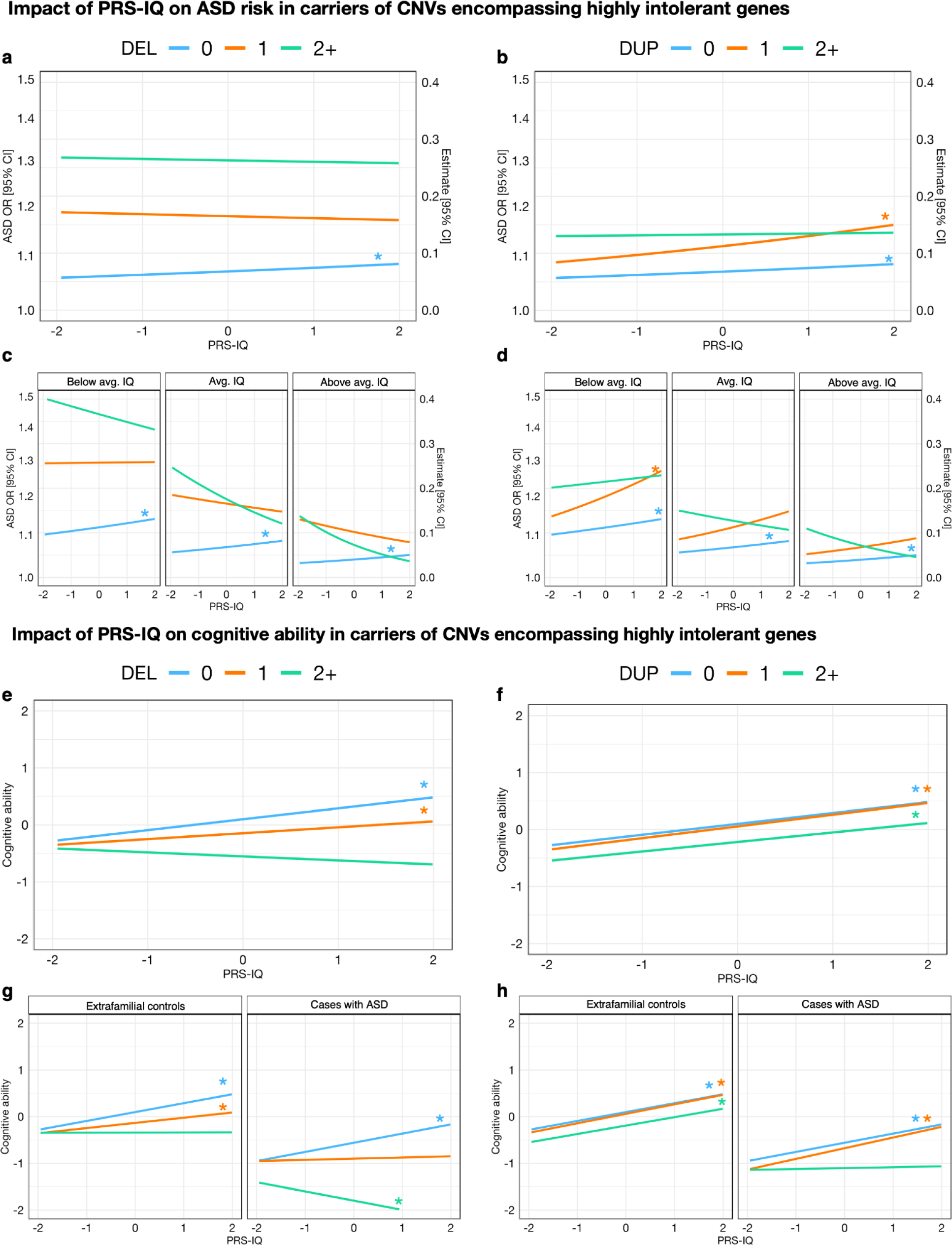
Deletions and duplications in highly intolerant genes modulate the impact of PRS-IQ on ASD risk and cognitive ability. Deconstructing the significant negative interaction between CNV burden and PRS-IQ that was identified in Figure 1. These interaction plots highlight the impact of highly intolerant CNVs in attenuating the effect of PRS-IQ on ASD risk and cognitive ability. **a,b)** Interaction plot showing the effect of PRS-IQ on ASD risk as moderated by the number of highly intolerant CNVs carried by each individual. PRS-IQ has a positive linear impact on ASD risk in individuals who do not carry any highly-intolerant deletions (n_cases_=8,170; n_controls_=168,499) or duplications (n_cases_=8,028; n_controls_=166,003). Carrying 1 deletion reduces the impact of PRS-IQ on ASD (n_cases_=162; n_controls_=1,049), and the negative effect of PRS-IQ on ASD risk is exacerbated in carriers of ≥2 deletions (n_cases_=94; n_controls_=256). Carrying highly intolerant duplications attenuates, but to a lesser extent compared to deletions, the effect of PRS-IQ on ASD risk (n_cases_=227; n_controls_=2512 carrying one duplication, and n_cases_=171; n_controls_=1,289 carrying ≥2 duplications). **c, d**) Adding cognitive ability as the second moderator reveals that these trends remain consistent, regardless of the scaled cognitive ability (1 standard deviation below the mean (-0.97), mean (0.067), or 1 standard deviation above the mean (1.10)) of the samples. **e, f**) Interaction plot showing the effect of PRS-IQ on cognitive ability as moderated by the number of highly intolerant CNVs carried by each subject. Except for individuals that carry ≥2 highly intolerant deletions, PRS-IQ has a positive linear impact on cognitive ability. However, carrying highly-intolerant deletions attenuated the impact of PRS-IQ on cognitive ability. The attenuation of PRS-IQ in the presence of highly intolerant deletions (**g**) and duplications (**h**) was more pronounced in cases with ASD compared to extrafamilial controls. Asterisks (*) denote statistically significant estimates of PRS-IQ, interacting with its moderator variable(s) (*P* value ≤ 0.05 following FDR adjustment for multiple corrections). While not all regression lines are statistically significant, these interaction plots should be interpreted as descriptive trends between the genetic risk factors and outcomes. See Table ST4 and Table ST5 for a detailed number of carriers and regression model results.

We then evaluated ASD risk and cognitive ability as a function of a three-way interaction between: 1–PRS-IQ as the predictor; 2–CNV carrier status as the moderator variable (separate terms for deletions and duplications), and either; 3–cognitive ability for the ASD risk model, or binary ASD diagnostic status for the cognitive ability model as the second moderator variable. The findings suggest that the impact of PRS-IQ on ASD risk and cognitive ability is attenuated in carriers of CNVs in highly intolerant genes, regardless of their cognitive ability (**Figures 3c, d**) and ASD case status (**Figures 3g, h**), respectively.

### The burden of CNVs and PRS-IQ converge to confer risk for ASD but diverge to confer risk for core and associated features and developmental history

We reasoned that since both CNVs and PRS-IQ increase ASD risk, they would affect at least one developmental traits (relevant to ASD) in the same direction. We therefore investigated the effect of intolerant CNVs and PRS-IQ on 19 cognitive, behavioural and developmental traits. However, this was not the case. Among these traits, 14 were significantly (FDR<0.05, 21 comparisons counting ASD and cognitive ability) impacted by both CNVs and PRS-IQ. No significant effects were identified for language regression, any regression, and the Social Responsiveness Scale. We observed that CNVs and PRS-IQ had diverging effects on all traits, including core and associated features of ASD, and developmental milestones (**Figure 4**a), similar to the diverging effects observed on cognitive ability but different from the converging effects observed on ASD. This is further demonstrated by hierarchical clustering (**figure 4 b**) based on effect sizes of all 3 classes of variants (deletions, duplications, PRS-IQ) which separates ASD itself from all other related phenotypes.

**Figure 4.**
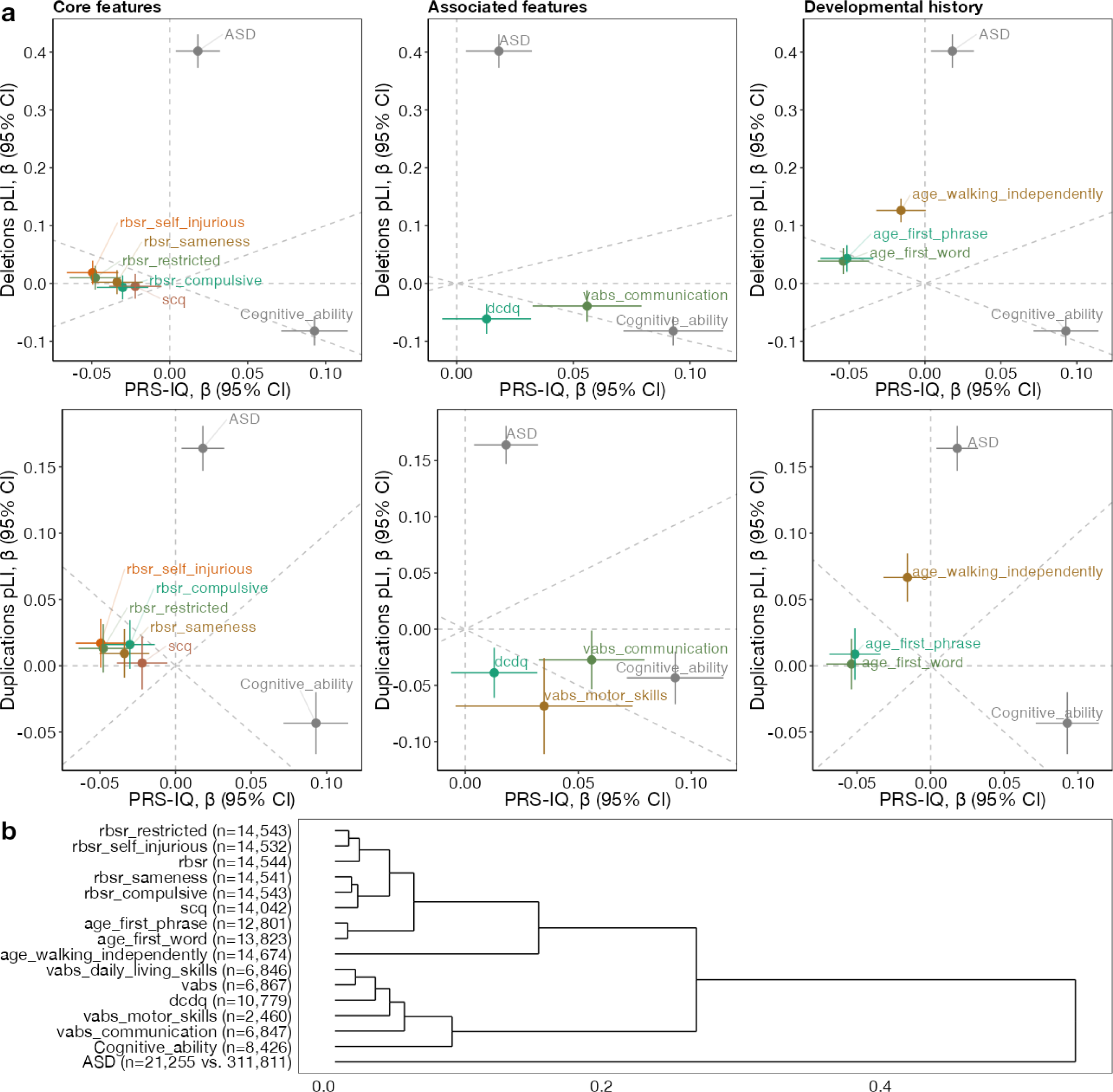
Genetic dissection of phenotypes among subjects with ASD. **a)** Comparing the directionality of PRS-IQ and deletions (upper plots), and duplications (lower plots). Effect sizes and their 95% confidence intervals on core ASD features, specifiers and associated features, and developmental history traits are displayed, along with ASD risk and cognitive ability. The burden of CNVs (deletions and duplications) and PRS-IQ only have convergent effects on ASD risk. **b)** Hierarchical clustering of phenotypes based on their association effect sizes with PRS-IQ, deletion burden and duplication burden. The number displayed reflects sample sizes. Only phenotypes with at least one significant effect (FDR<0.05) are displayed within each plot. See section “Phenotypic measures” for references to abbreviations.

## DISCUSSION

This study dissects the paradoxical effects of intolerant CNVs and PRS-IQ that both increase ASD risk while impacting cognitive ability in opposing directions. We show that the effects of intolerant CNVs and PRS-IQ increase the risk for ASD, even after adjusting for their effects on cognitive ability. This suggests that cognitive ability may not be the dimension underlying ASD risk conferred by both of these classes of genetic variants. To further demonstrate that decreased cognitive ability may not be the factor underlying liability for ASD, we show that the bottom decile of PRS-IQ is protective for ASD after adjusting for its effects on cognition. Combining both classes of variants (CNVs and PRS) suggests a negative interaction, such that the impact of PRS-IQ on ASD risk and cognitive ability is attenuated in carriers of intolerant CNVs (see **Figure 5** for a summary of the results). However, larger sample cohorts and independent replications are required to confirm this observation. Finally – unlike their impact on ASD risk, we find that CNVs and PRS-IQ have opposing effects on developmental phenotypes among cases with ASD.

**Figure 5.**
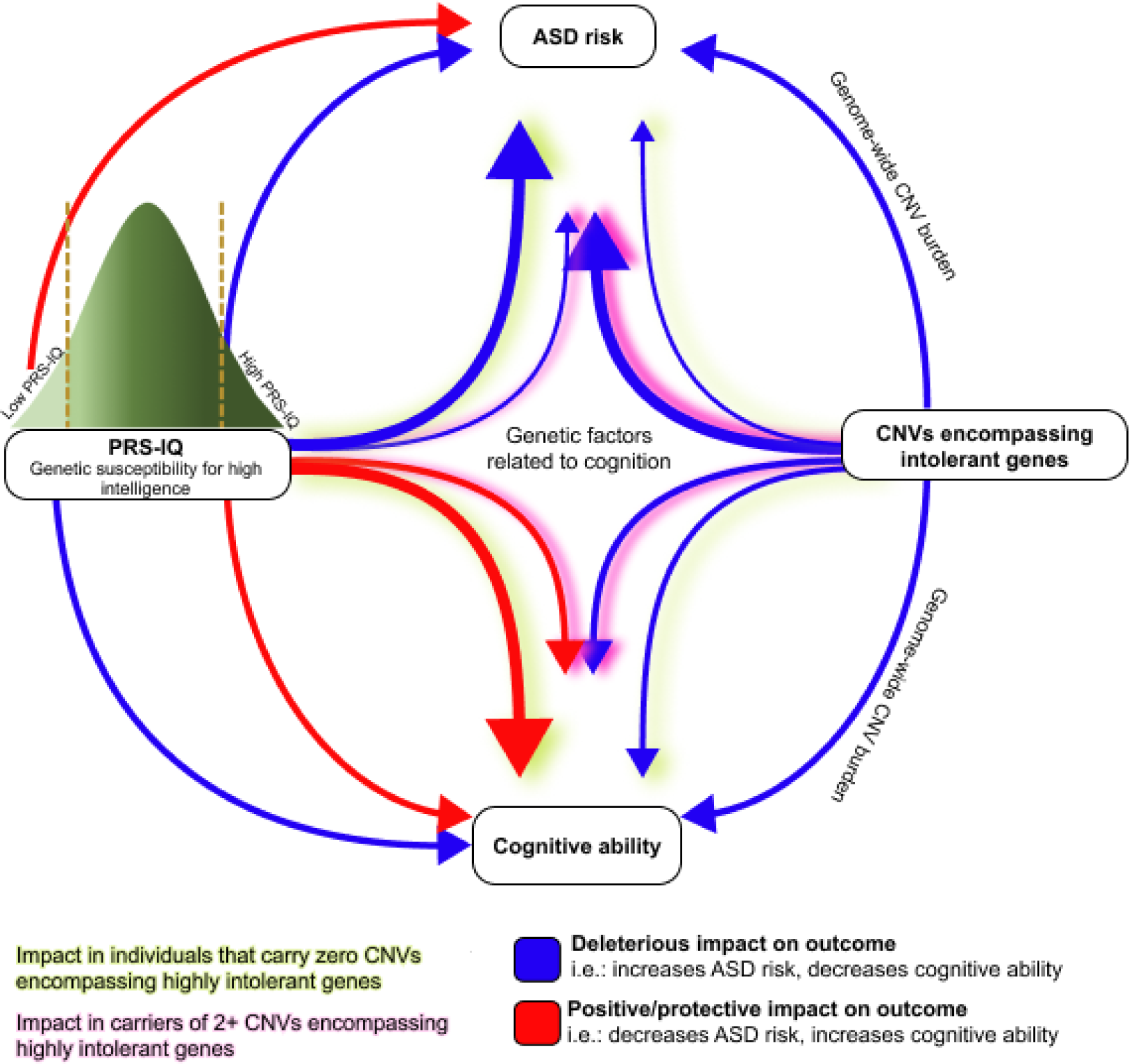
Summary of the impact of cognition-related genetic factors on ASD risk and cognitive ability. Representation of the main findings from this study. Of note, the impact of cognition-related genetic factors on ASD risk remains unchanged, even after adjusting for cognitive ability in the models. The thickness of the arrows denotes the relative impact of the variant class on the outcome.

Low cognitive ability is a major feature of ASD and is associated with greater autistic impairments^44^. We find that intolerant deletions and duplications follow this pattern, such that they decrease cognitive ability and increase ASD risk. Conversely, we show that common genetic risk factors that are similarly associated with low cognitive ability (i.e.: bottom PRS-IQ decile) are protective against a diagnosis of ASD. In fact, these findings suggest that intolerant CNVs and PRS-IQ may modulate cognitive ability and ASD risk through distinct mechanisms. While this phenomenon may be driven by the role of cognition in the evolutionary mechanisms of ASD and the shared features between ASD and high IQ (i.e.: large brain size, fast brain growth, enhanced synaptic functions, increased attentional focus, positive assortative mating)^45–47^, further studies are warranted to delineate the complex relationship between these factors.

We identified a significant negative interaction between these genetic risk factors, implying that the impact of PRS-IQ on ASD risk is attenuated in subjects with a high burden of CNVs, irrespective of their level of cognitive ability. However, the significance and effect size of this interaction is weak suggesting that most of the CNV and PRS-IQ effects are additive. Similar additive effects between CNV and PRS have been previously observed in schizophrenia ^48, 49^. In our study, however, we show that such an additive effect extends to two classes of variants with opposing effects on the same trait (cognitive ability), further highlighting the fact that ASD risk is independent of the effects of variants on cognitive ability. These findings support the liability threshold model for ASD, in which common variants and CNVs act additively to confer risk for the disorder. We also identify a similar relationship between PRS-IQ and CNVs in modulating cognitive ability such that deleterious CNVs reduce the positive impact of PRS-IQ on cognitive ability. Interestingly, our results suggest that individuals with ASD are more susceptible to the deleterious impact of intolerant CNVs on cognitive ability compared to the general population. Our findings suggest that individuals with a genetic risk for low (i.e.: high CNV burden, deleterious CNVs, or bottom PRS-IQ deciles) or high (i.e.: top PRS-IQ deciles) cognitive ability have a substantial risk for ASD, irrespective of their level of cognitive ability. Interestingly, we find that cases with ASD carry substantial genetic risk for either low or high cognitive ability, but not both.

Beyond the opposing effects on cognitive ability, this study shows that these two classes of genetic risk for ASD systematically show opposing effects on all developmental milestones and dimensional traits, including core features of ASD. This highlights the fact that biological factors which increase ASD risk may not necessarily map onto core ASD features.

## LIMITATIONS

This is the first large-scale study that controls for cognitive ability when modelling the impact of cognition-related genetic factors on ASD risk. Yet, it is important to consider that the available IQ measures only serve as a proxy to capture cognitive ability. Furthermore, the dimension of cognitive ability used in the study consists of diverse cognitive measures across cases and controls of different age groups. Moreover, impairments in social communication may limit the performance of traditional IQ measurements and underestimate the cognitive abilities of individuals with ASD^50–52^. While we identified a significant negative interaction between CNVs encompassing intolerant genes and PRS-IQ in ASD risk and cognitive ability, larger sample sizes are needed to confirm this interaction. This is an intrinsic limitation in studying the integrated effects of rare and common variants: even though the present study assembles some of the largest community-based cohorts as well as ASD cohorts presently available, the sample sizes for carriers of rare CNVs remain small (highly intolerant genes deletions or duplications are present in <5% of individuals globally). Thus, the study of the combined effects of these variants with PRSs is subject to class imbalance. As such, the interaction plots we present should be interpreted with caution – solely as an exploratory analysis of the trends between these genetic factors – given that the models are trained on a smaller subset of the sample. While these findings capture the interaction between CNVs and PRS-IQ in modulating ASD risk and cognitive ability, we lack sufficient statistical power to robustly ascertain this effect in each analysis group.

## CONCLUSIONS

Rare CNVs and PRS-IQ increase the risk of ASD independently of their opposing effects on cognitive ability. Altogether, this study supports the complex interplay between these two classes of genomic variants on ASD risk, cognitive ability, and ASD-associated features. Our findings suggest that these genetic factors related to cognitive ability may be a key element in ASD risk. Indeed, there remains insufficient evidence for ASD-specific genes^5^, which puts the relevance of searching for ASD-specific – rather than cognition-related – genetic risk factors into question. As such, we posit that cognitive ability may have been a red herring in the interpretation of genetic studies in ASD. Indeed, searching for ASD-associated genomic variants that do not influence cognitive ability may not be necessary to advance our understanding of the mechanisms underlying ASD risk. Finally, we also show that the way genetic variants influence dimensional traits may not fully inform on the mechanisms underlying their liability for ASD.

## Data Availability

All data produced in the present study are available upon reasonable request to the authors.

## ETHICS

All individuals or their legal representatives gave their written informed consent. All procedures were carried out according to the Declaration of Helsinki. Regarding the access to the cohorts and databases, approvals were obtained from the involved institutions. All data were deidentified.

Ethical approval for the Generation Scotland study was obtained from the Tayside Committee on Medical Research Ethics (on behalf of the National Health Service.

LBC1936 ethical approval was obtained from the Multicentre Research Ethics Committee for Scotland (MREC/01/0/56) and the Lothian Research Ethics Committee (LREC/2003/2/29).

## ACKNOWLEDGEMENTS

We are grateful to all the families who took part, the general practitioners and the Scottish School of Primary Care for their help in recruiting them, and the whole Generation Scotland team, which includes interviewers, computer and laboratory technicians, clerical workers, research scientists, volunteers, managers, receptionists, healthcare assistants and nurses.

The authors thank all LBC study participants and research team members who have contributed, and continue to contribute, to ongoing studies.

