## Supplemental Figures for "Copy-number variants and polygenic risk for intelligence confer risk for autism spectrum disorder irrespective of their effects on cognitive ability"

### Supplementary Figures

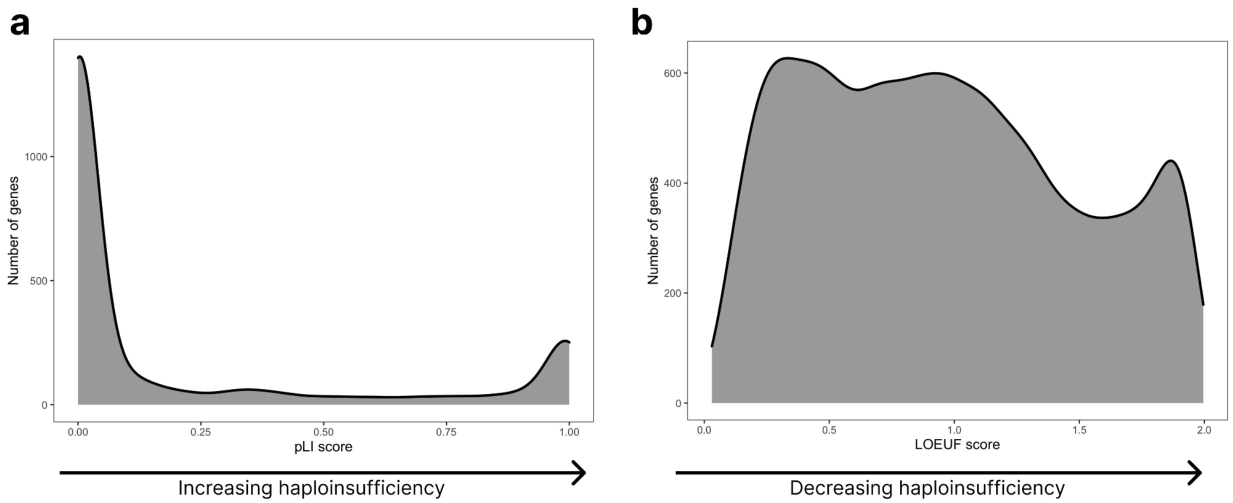


**Figure S1. Distribution of haploinsufficiency scores across 19,197 genes.**
**a**) The distribution of the probability of being loss-of-function intolerant (pLI) annotation. The bimodal distribution better captures the genome-wide burden of CNV risk across each individual. b) The distribution of the loss-of-function observed/expected upper bound fraction (LOEUF) annotation. The continuous metric facilitates the stratification of individuals according to their carrier status of CNVs encompassing haploinsufficient genes,

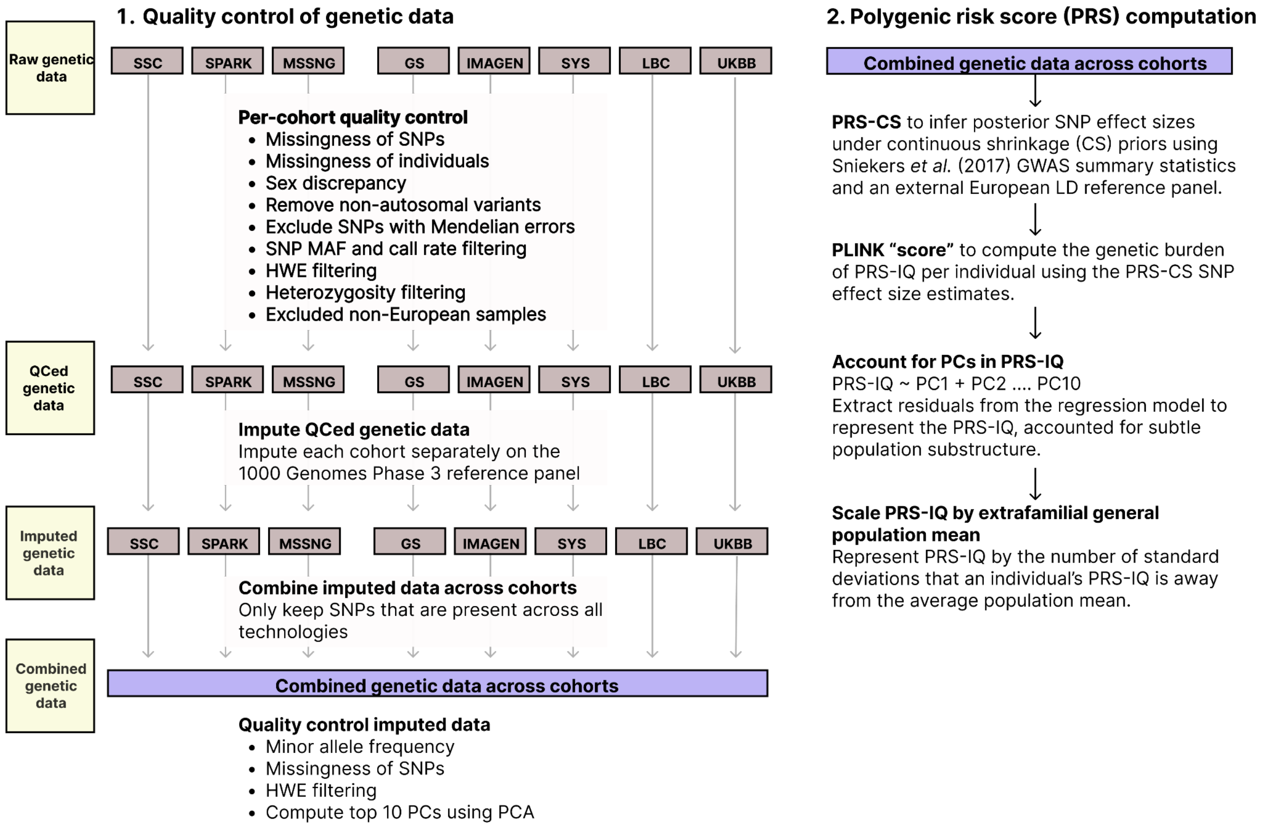


**Figure S2. Methodological pipeline to compute and combine PRS-IQ data across eight cohorts.** Description of each analysis step used to compute PRS-IQ across eight cohorts. To ensure that the PRS-IQ data across cohorts were comparable within each analysis group (cases with ASD, intrafamilial controls, and extrafamilial controls), we performed a one-way ANOVA and pairwise t.test (see: Tables ST2 and ST3). There was no significant difference in the mean PRS-IQ across cohorts following the final analysis step.

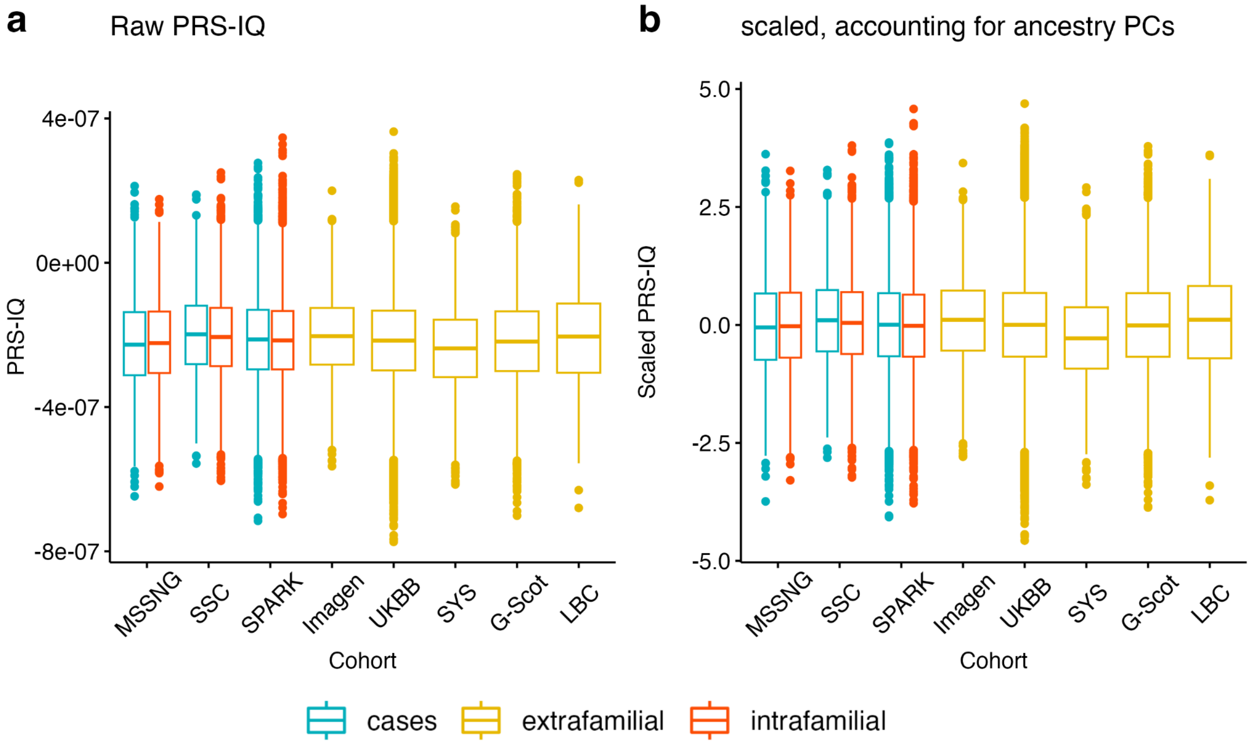


**Figure S3. Distribution of PRS-IQ across cohorts in each analysis group.**
The distribution of PRS-IQ across the different PRS computation steps: the "raw" PRS-IQ across all European samples (**a**), and; the scaled residuals after modelling PRS-IQ as a function of the top 20 ancestry PCs (**b**). We combined the imputed data such that only the variants that were present across all genotyping and sequencing technologies were retained. This step was important to reduce PRS-IQ biases due to batch effects. There was no difference in PRS-IQ between intrafamilial and extrafamilial controls (Figure 2c), which were derived from distinct cohorts. This validated our approach and findings that compared PRS-IQ between cases with ASD and extrafamilial controls.

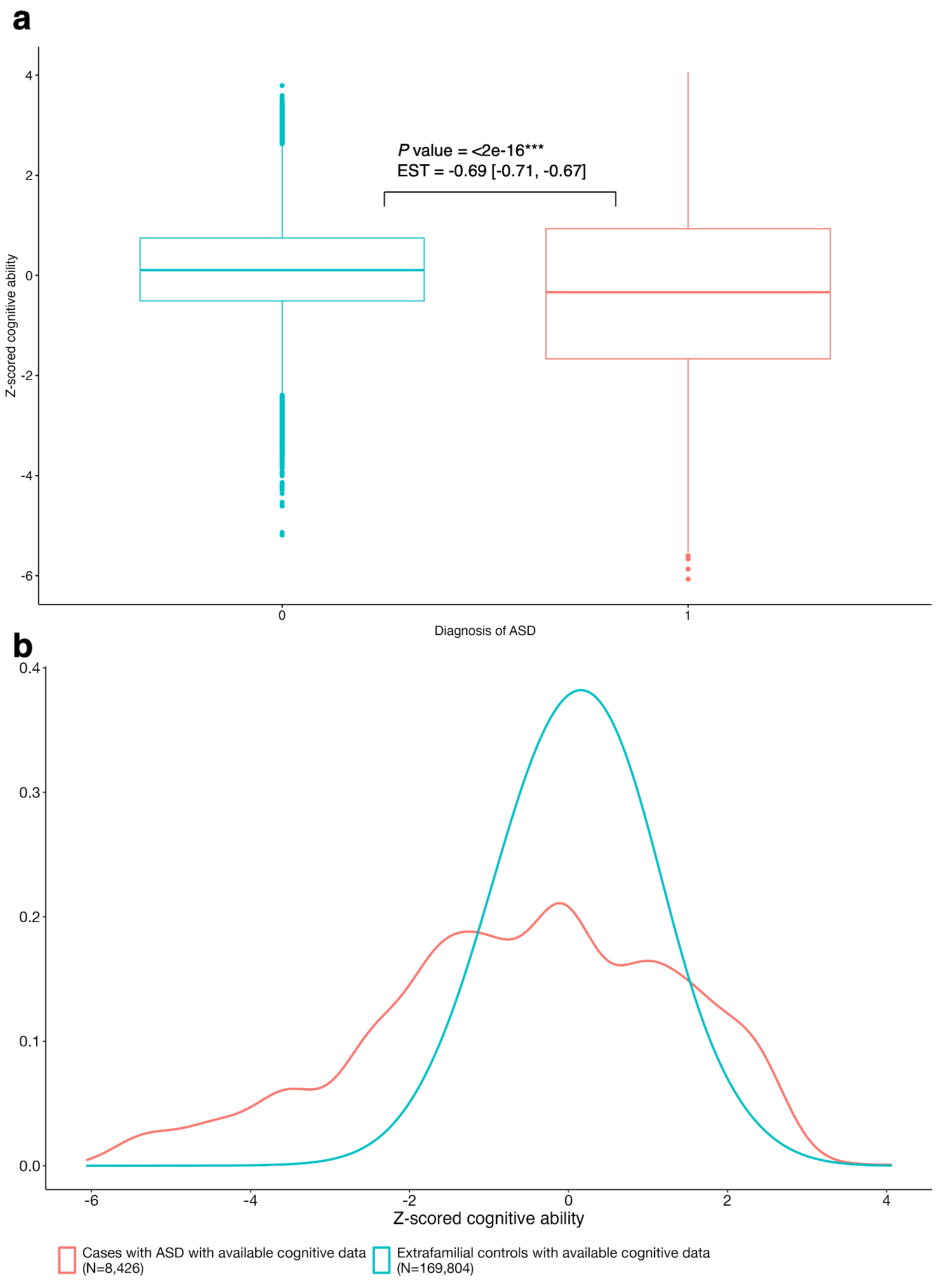


**Figure S4. Distribution of scaled cognitive ability.**

**a**) On average, the mean cognitive ability of subjects with ASD was 0.69 units lower than extrafamilial controls. *P* value was the result of a linear regression using ASD status as predictor, and cognitive ability as the outcome. The model included sex as a covariate. **b**) Density plot representing the distribution of scaled cognitive measures across cases with ASD (N=8,426) and extrafamilial controls (N=169,804) for whom cognitive ability data was available. The density of the curve was smoothed using a bandwidth selector of 0.4 for the Gaussian kernel density estimation.

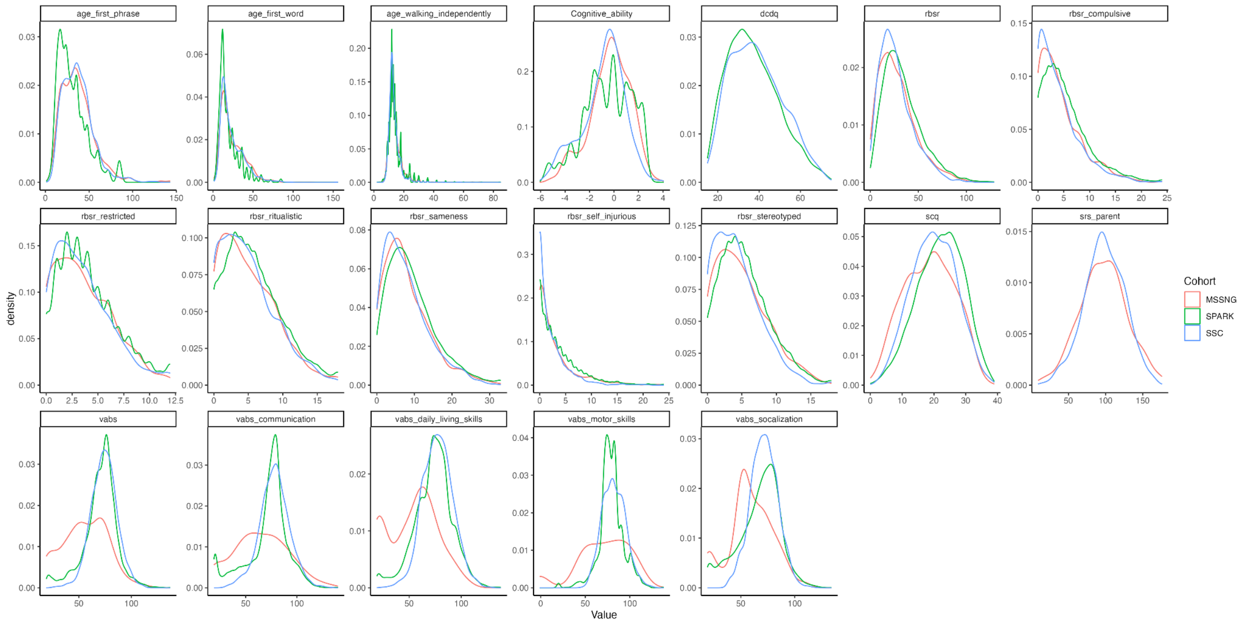


**Figure S5. Distribution of raw continuous phenotypic measures among individuals with ASD, for the three ASD cohorts.**
For a detailed description and abbreviation of the traits included in the study, see "Phenotypic measures" in Methods section of the manuscript.

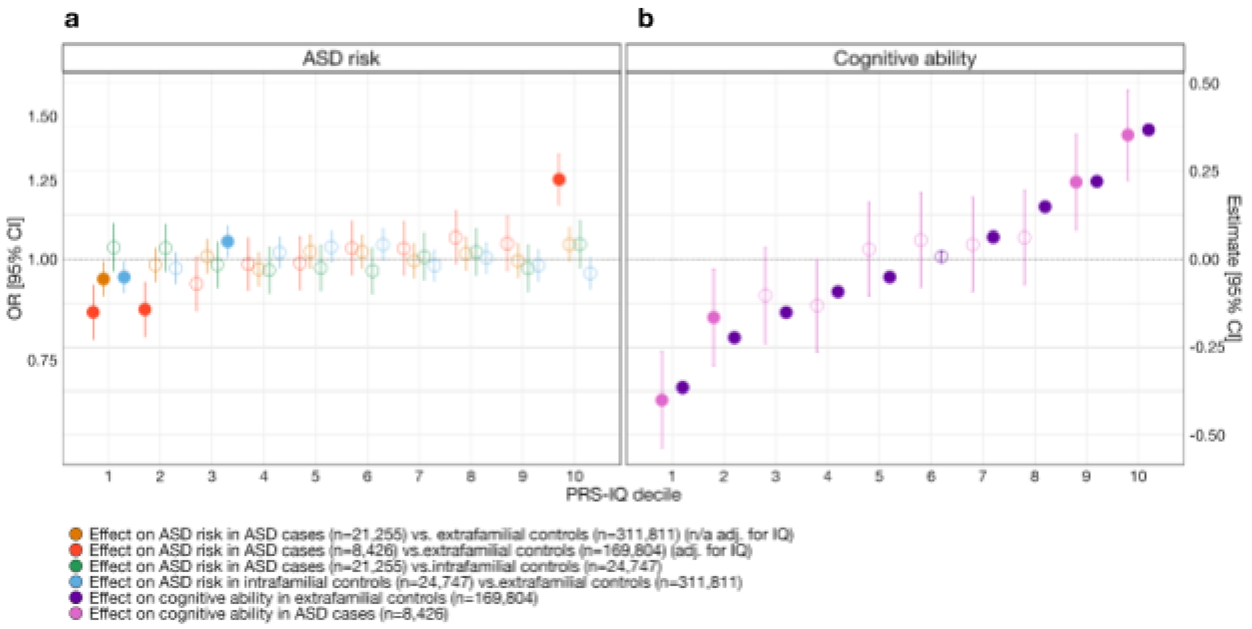


**Figure S6. The effect of PRS-IQ deciles on ASD risk and cognitive ability across all analysis groups.** The estimate and 95% CI of each PRS-IQ decile on ASD risk and cognitive ability. Each regression accounted for the individual-level burden of deletions, duplications, and – when available – cognitive ability of individuals included in the model. **a**) Even after adjusting for the effects of cognitive ability (red points), a high PRS-IQ (10th decile) increases the risk for ASD, while a low PRS-IQ (1st, 2nd deciles) decreases the risk for ASD. Cases with ASD in the lowest PRS-IQ decile also have a significantly lower risk for ASD than their unaffected family members. **b**) A PRS-IQ below and above the 6th decile significantly decreases and increases cognitive ability in the general population, respectively. The effect of PRS-IQ on cognitive ability is the same in cases with ASD and in the general population (extrafamilial controls). Filled-in points represent statistically significant terms (*P* value ≤ 0.05 following FDR adjustment for multiple corrections). For detailed model results, see **Table ST6**.
